# Hypertension and renin-angiotensin system blockers are not associated with expression of Angiotensin Converting Enzyme 2 (ACE2) in the kidney

**DOI:** 10.1101/2020.05.19.20106781

**Authors:** Xiao Jiang, James Eales, David Scannali, Alicja Nazgiewicz, Priscilla Prestes, Michelle Maier, Matthew Denniff, Xiaoguang Xu, Sushant Saluja, Eddie Cano-Gamez, Wojciech Wystrychowski, Monika Szulinska, Andrzej Antczak, Sean Byars, Maciej Glyda, Robert Król, Joanna Zywiec, Ewa Zukowska-Szczechowska, Louise M. Burrell, Adrian S. Woolf, Adam Greenstein, Pawel Bogdanski, Bernard Keavney, Andrew P. Morris, Anthony Heagerty, Bryan Williams, Stephen B. Harrap, Gosia Trynka, Nilesh J. Samani, Tomasz J. Guzik, Fadi J. Charchar, Maciej Tomaszewski

## Abstract

Angiotensin converting enzyme 2 (ACE2) is the cellular entry point for severe acute respiratory syndrome coronavirus (SARS-CoV-2) – the cause of COVID-19 disease. It has been hypothesized that use of renin-angiotensin system (RAS) inhibiting medications in patients with hypertension, increases the expression of ACE2 and thereby increases the risk of COVID-19 infection and severe outcomes or death. However, the effect of RAS-inhibition on ACE2 expression in human tissues of key relevance to blood pressure regulation and COVID-19 infection has not previously been reported.

We examined how hypertension, its major metabolic co-phenotypes and antihypertensive medications relate to ACE2 renal expression using information from up to 436 patients whose kidney transcriptomes were characterised by RNA-sequencing. We further validated some of the key observations in other human tissues and/or a controlled experimental model. Our data reveal increasing expression of ACE2 with age in both human lungs and the kidney. We show no association between renal expression of ACE2 and either hypertension or common types of RAS inhibiting drugs. We demonstrate that renal abundance of ACE2 is positively associated with a biochemical index of kidney function and show a strong enrichment for genes responsible for kidney health and disease in ACE2 co-expression analysis.

Collectively, our data indicate that neither hypertension nor antihypertensive treatment are likely to alter individual risk of SARS-CoV-2 infection or influence clinical outcomes in COVID-19 through changes of ACE2 expression. Our data further suggest that in the absence of SARS-CoV-2 infection, kidney ACE2 is most likely nephro-protective but the age-related increase in its expression within lungs and kidneys may be relevant to the risk of SARS-CoV-2 infection.

## INTRODUCTION

Severe acute respiratory syndrome coronavirus 2 (SARS-CoV-2) is responsible for the coronavirus disease 2019 (COVID-19) – the recent viral pandemic with high mortality rates and overwhelming burden on the healthcare systems globally. The virus gains entry into human host cells upon binding to angiotensin converting enzyme 2 (ACE2) – a molecule operating both as the main transmembrane receptor for the virus^1,2^ and a component of renin-angiotensin system (RAS) – the key blood pressure (BP) regulating cascade^3,4^. Interestingly, elevated blood pressure (hypertension) has been implicated as a main co-morbidity and a potential risk factor for more severe clinical outcomes of COVID-19^5,6^, and speculation mounted that this may be due to commonly prescribed antihypertensive medications targeting RAS [i.e. angiotensin converting enzyme inhibitors (ACE-I) or angiotensin II type 1 receptor (AT1) antagonists (ARB)]. Indeed, RAS blockers have been hypothesised to enhance SARS-CoV-2 entry into the host cells and/or promote the organ damage in patients with COVID-19 as a result of ACE2 up-regulation^7–10^. However, evidence for a direct effect of hypertension or RAS blockers on ACE2 expression in human tissues has remained elusive, largely because of the paucity of large gene expression datasets with matching clinical information. As the inhibition of the ACE2-SARS-CoV-2 interaction gains traction as a potential treatment strategy for COVID-19^11^, an urgent understanding of how the key COVID-19 comorbidities alter ACE2 expression in human tissues is necessary to gain new mechanistic and therapeutic insights.

Herein, we selected the human kidney as a tissue of key importance to BP regulation, RAS and COVID-19 and examined how hypertension (as well as its major metabolic co-phenotypes) related to ACE2 expression^7,12–15^. We further explored if RAS blockers were associated with changes in ACE2 expression taking advantage of information on antihypertensive treatment in patients whose kidney transcriptomes were characterised by RNA-sequencing. Finally, we validated some of the key observations in other human tissues and a controlled experimental model using kidneys from spontaneously hypertensive rats (SHRs) treated with ACE-I and ARB.

## METHODS

### Human kidney tissue collections

We used 436 human kidney samples collected from patients recruited into five studies [TRANScriptome of renaL humAn TissuE (TRANSLATE) study^16–21^, its extension – TRANSLATE-T study^18^, moleculAr analysis of human kiDney-Manchester renal tIssue pRojEct (ADMIRE)^20^, molecular analysis of mechanisms Regulating gene Expression in Post-ischAemic Injury to Renal allograft (REPAIR)^20^ and Renal gEne expreSsion and PredispOsition to cardiovascular and kidNey Disease (RESPOND)^20^] in our discovery resource. In TRANSLATE, ADMIRE and RESPOND studies, the samples were collected from patients with unilateral kidney cancer – the specimen was taken from unaffected by cancer part of the kidney immediately after elective nephrectomy^16-21^. TRANSLATE-T and REPAIR collected pre-implantation kidney biopsies^18^ from deceased kidney donors prior to the organ transplantation^18,20^. The secured tissue samples from all the studies were immersed immediately in RNAlater or snap-frozen for the purpose of further molecular analysis. We used an additional cohort of 98 individuals from the NIH-supported Tissue Cancer Genome Atlas (TCGA)^22,23^ as a replication resource for some of our kidney gene expression analyses. We only used samples collected as “companion normal tissue specimen” – those taken from healthy (unaffected by cancer) part of the kidney after the surgery^23^, in line with the tissue collection strategy used by TRANSLATE study. All patients were of white-European ethnicity.

### Phenotypes

For the purpose of this project we extracted the relevant demographic and clinical information (age, sex, body weight index, height, hypertension, diabetes, renal function) from the discovery projects’ databases. We calculated body mass index by dividing weight (in kg) by height^2^ (in m). In TRANSLATE and RESPOND studies, hypertension was defined as BP values ≥140/90 mmHg (measured on at least two separate occasions) and/or being on pharmacological antihypertensive treatment, as reported elsewhere^17^. Diabetes was defined as either self-reported history of diabetes and/or being on hypoglycaemic medications^19^. In ADMIRE, TRANSLATE-T and REPAIR studies, information on history of hypertension and diabetes was recorded by the respective recruitment teams based on the available hospital documentation. All hypertensive patients had information on whether they were managed using BP lowering medications. A total of 72.4% of hypertensive patients on pharmacological antihypertensive treatment had further detailed information on the prescribed antihypertensive medications. In these patients we allocated each of the prescribed antihypertensive medication into one of the following antihypertensive classes: ACE-I, ARB, beta-blockers (BB), calcium channel antagonists (CCA), diuretics (DRT) or others. Serum levels of creatinine were measured by modified kinetic Jaffe method (calibrated to an IDMS reference measurement procedure) in 310 individuals. In calculating estimated glomerular filtration rate (eGFR) we used CKD-EPI equation^24^, as reported before^19^. Only basic demographic information (age, sex) were available for patients recruited in the kidney replication resource (TCGA)^19^.

### Genotyping and genetic principal components

In the discovery resource, kidney DNA was extracted using Qiagen DNeasyBlood and Tissue Kit, and then hybridised to the Infinium® HumanCoreExome-24 beadchip array, as reported before^18,20,21^. In TCGA, DNA was extracted from blood samples using QiAAmp Blood Midi Kit (CGARN, 2016) and hybridised with probes on the Affymetrix SNP 6.0 array^23^. We applied the same quality control filters to genotyped variants and individuals in all data sets. In brief, we excluded autosomal genotyped variants based on the following criteria: genotyping rate <95%, Hardy-Weinberg equilibrium (HWE) P<1×10^−3^, minor allele frequency (MAF)<5%, position duplicated variants^18,20^. Prior to further analyses we removed individuals with: genotyping rate <95%, heterozygosity rate outside ±3 standard deviations from the mean, cryptic relatedness to other individuals, inconsistent sex information and non-white-European genetic ancestry^18,20^. Genotype principal components (PCs) were calculated using PLINK v1.90b6.2^25^. In the calculation we firstly removed ambiguous genetic variants (C/G or A/T) with MAF>0.4. Genetic variants in 24 high linkage disequilibrium (LD) regions were removed thereafter. We then used “--indep-pairwise” in PLINK to further prune variants in LD setting window size of 1000bp, step size of 50 variants and r^2^ cut-off threshold as 0.05. We calculated the genotype PCA for each of the discovery and replication resource, using “--pca” in PLINK.

### Kidney transcriptome profiling

We used RNeasy kits (Qiagen) to extract RNA from kidney tissue in 75% of samples in our discovery resource. The remaining samples were subjected to an RNA extraction using miRNeasy Mini Kit (Qiagen). In the replication resource, kidney RNA was extracted from snap-frozen samples using a modification of the DNA/RNA AllPrep Kit (Qiagen), as reported before (https://brd.nci.nih.gov/brd/sop/show/1450).

Sequencing libraries were generated from 1 μg of extracted RNA using Illumina TruSeq poly-A protocol. Libraries were then sequenced using either 100 bp reads (on an Illumina HiSeq 2000) or 75 bp paired-end reads (on an Illumina NextSeq or HiSeq 4000) producing an average of 32 million paired reads and 5.5 Gb per sample. The base calling and sequence quality were evaluated using FastQC^26^ on all sequenced libraries. We used Kallisto to quantify the expression of genes at a transcript level [in transcripts per million (TPM)]^27^ and then summed them to generate gene-level expression values, as reported elsewhere^20^. For the purpose of our downstream computational analyses we selected genes with the expression >TPM of 0.1 and the read count ≥6 (in at least 20% of kidney samples, within each study)^20^. All sequenced samples were examined using several quality control filters including: number of total reads (>10million reads), D-statistic test (a normalised measure of within tissue sample inter-correlation, D≤5), sex compatibility check (consistency between the reported sex and expression of sex-specific genes, based on XIST and expression of RPS4Y1, KDM5D, DDX3Y, EIF1AY and USP9Y), verification of sample code based on comparing DNA base calls obtained from RNA-sequencing using GATK and DNA genotype calls and visual inspection of principal component plots of processed TPM data.

After applying these quality control filters, 21,203 renal genes (common for all studies in the discovery and replication dataset) were retained for the downstream analyses. Prior to the downstream analyses, gene expression data that passed the quality control underwent normalisation. The expression values underwent first log-transformation [natural logarithm of TPM (plus an offset of 1)] followed by quantile normalisation (http://bioconductor.org/packages/release/bioc/html/aroma.light.html), as reported elsewhere^20^. The quantile-normalised data were then standardised using rank-based inverse normal transformation^18^.

### Kidney single-cell RNA-sequencing analysis

Single-cell RNA-sequencing data from the human adult kidney was obtained from Young et al.^28^ and re-analysed^20^. Briefly, we used data from 41,778 normal kidney cells, combined this with literature-curated markers to annotate cell types and used Seurat^29^ to determine cell type marker genes and a two-dimensional t-distributed stochastic neighbour embedding (t-SNE) of the expression data^20^. We identified 23 distinct cell clusters. These were then grouped into 8 renal cell types, 5 immune cell types and 1 unidentified by their respective expression of canonical cell type markers. The 8 renal cell types were then grouped and visualised by their localisation to each nephron segment.

### Kidney ACE2 co-expression analysis

The ACE2 co-expression analysis employed multivariate regression to determine the association between each expressed kidney gene and ACE2; models were adjusted for age, sex, diabetes, hypertension, three genetic PCs and surrogate variables automatically inferred by the “sva” R package in line with Tukiainen et al.^30^. This analytical pipeline was applied to the discovery and replication data sets separately. The crude P-values were then adjusted by permutation-based resampling. The crude t-statistics were scaled by the mean and standard deviation of t-statistics from 1,000 permutations of the data, then the P-values were re-calculated and the FDR-based adjustment for multiple testing was applied. A gene was considered as co-expressed with ACE2 if the following criteria were met: (i) FDR <0.05 in the discovery data, (ii) FDR <0.05 in the replication data and (iii) consistent direction of association with ACE2 in discovery and replication.

Gene set overrepresentation for REACTOME pathways and gene ontology cellular compartment annotation was performed by the PANTHER classification system^31^ employing Fisher’s exact test. Official gene symbols for all ACE2 co-expressed genes were uploaded as an input list to the PANTHER analysis and were compared against all human genes. The kidney disease and BP gene sets were built from a combination of GWAS catalog (https://www.ebi.ac.uk/gwas/) association data for phenotypes “Blood pressure” (EF0_0004325) and “Kidney disease” (EF0_0003086) and manually curated genetic associations from the GWAS literature. Their enrichment among ACE2-coexpressed genes was examined using Fisher’s exact test and all human genes as a comparator.

### Renal expression of ACE2 – association analyses

We used ordinary least square linear regression models to examine associations between normalised renal expression of ACE2 [or sex-related “control” genes (XIST, RPS4Y1)] as dependent variables with each of the selected demographic/clinical variables – age, sex, body mass index (BMI), hypertension, diabetes and eGFR as independent parameters; separately in the discovery and the replication resource (where available). All the regression models were adjusted for (i) first three genotype PCs – to adjust for the population structure, (ii) a set of demographic (age, sex) and clinical (i.e. BMI, diabetes; where appropriate) parameters – to minimise their potentially confounding effects on the dependent variable and (iii) surrogate variables (SVs) – to remove the batch effects such as those generated by variation in recruitment centres, DNA and RNA processing protocols, and times of sequencing^32^. The SVs also remove the variation from unmeasured and/or technical cofounders, including but not limited to the sources of tissues, effects of differences in cell types^33^. The SVs were calculated for each model separately. The optimised number of surrogate variables for each model was determined by the “sva” package^32^, in line with previous studies^30,34^. The results from the analysis of the association between renal expression of ACE2 and sex in the discovery and replication resources were combined using an inverse-variance fixed-effect meta-analysis^35^.

The analysis of the association between renal expression of ACE2 and each of the antihypertensive class was adjusted for age, sex, BMI, diabetes, first three genotype PCs and SVs, and further corrected for the inter-correlation between the drug classes (using “gee” R package). The absolute normalised and adjusted differences in renal ACE2 between those who were taking a specific class of antihypertensive medications versus those who were not on any medication were converted into fold differences using the exponential values of the corresponding estimated coefficients. The correction for multiple testing was calculated using “qvalue” package^36,37^ in R.

### Expression of ACE2 in other human tissues – Genotype-Tissue Expression project (GTEx)

We also explored the gene expression information from Genotype-Tissue Expression project (GTEx) – an NIH-sponsored publicly available resource^38^. We extracted information on normalised ACE2 expression from the transcriptome of the lung as well as the following human tissues of relevance to hypertension/BP regulation: adrenal gland, visceral adipose tissue, subcutaneous adipose tissue, left ventricle, atrial appendage, tibial artery, coronary artery and aorta^38^. The number of post-mortem donors who contributed transcriptomic information for these tissues was 515, 233, 469, 581, 386, 372, 584, 213 and 387 (respectively). Analysis of association between ACE2 expression and age as well as sex were completed separately in each tissue using a multivariate regression model and the “Limma” R package^39^. All models were adjusted for age, sex (as appropriate), BMI, sample ischaemic time, death classification on the Hardy scale^30^, 3 genetic PCs and a tissue-specific number of SVs inferred by the “sva” R package. The nominal P-values were further adjusted by FDR.

### Kidney expression of ACE2 and eGFR – replication analysis in Nephroseq

To replicate the association between kidney ACE2 expression and eGFR, we examined renal gene expression data sets curated by Nephroseq (www.nephroseq.org). Included were gene expression studies on human renal tissue (tubulointerstitium) with a minimum of 10 informative individuals in the analyses. Five datasets from four studies [including European Renal cDNA Bank (ERCB), Ju et al. – discovery and validation^40^, Sampson et al.^41^, Woroniecka et al.^42^] were eligible; in ERCB transcriptome profiling was conducted using RNA-sequencing, in the other studies – by microarrays. For each of the studies we computed Pearson’s correlation coefficient for renal tubulointerstitial expression of ACE2 against eGFR. To determine the overall correlation between renal ACE2 expression and eGFR we then combined the information from all studies with the Olkin-Pratt fixed-effect meta-analytical approach (using the R package Metacor). Heterogeneity was examined using Cochran’s Q test.

### Kidney expression of ACE2 in rats treated with perindopril and losartan

To examine ACE2 expression in response to losartan and perindopril, we first sourced 6-week old inbred male spontaneously hypertensive rats (SHR) from the Animal Resources Centre (Canning Vale, Western Australia). At 10 weeks old, SHR were implanted with minipumps (Alzet model 2004, Durect Corp Cupertino, California) for infusions of either the vehicle (normal saline), losartan (7.5 mg/kg/d) or perindopril (1 mg/kg/d). SHR received infusions for 4 weeks, after which treatment ceased at 14 weeks old. SHR were sacrificed at 14 weeks old (n=4-10) under general anaesthetic with isoflurane and ketamine. The kidneys were removed, decapsulated and dissected; renal cortices were subsequently submerged in RNAlater stabilisation solution (Thermo Fisher Scientific), frozen in liquid nitrogen and stored at −80°C. RNA was later extracted from the isolated cortices using the mirVana™ miRNA isolation kit (Thermo Fisher Scientific) and quantified using the NanoDrop 2000 (Thermo Fisher Scientific). To account for technical variation, gene expression of ACE2 was normalised against housekeeping gene (GAPDH) using quantitative real-time PCR. We then performed one-way ANOVA and unpaired student t-tests on ΔCt values using GraphPad Prism 8 to assess variability between groups and calculated fold change from ΔCt values equalling to 2-ΔΔCt with ΔΔCt = ΔCt(sample) – ΔCt(control).

### Bioethics

The human studies adhered to the Declaration of Helsinki and were approved/ratified by the appropriate Bioethics Committee of the Medical University of Silesia (Katowice, Poland), Bioethics Committee of Karol Marcinkowski Medical University (Poznan, Poland), Ethics Committee of University of Leicester (Leicester, UK) and the University of Manchester Research Ethics Committee (Manchester, UK). Informed written consents were obtained from all individuals – for the deceased donors from TRANSLATE-T, the consent was obtained from the members of the family. All animal experiments were approved by the University of Melbourne Animal Ethics Committee and were conducted in accordance with the Australian Code of Practice for the Care and Use of Animals for Scientific Purposes.

## RESULTS

### Renal expression of ACE2 – insights from kidney transcriptome profiling, co-expression analysis and single-cell experiments

We first processed RNA-sequencing-derived gene expression profiles of 436 kidneys from our discovery resource (*TableS1*) and uncovered 21,203 kidney genes. The key RAS genes showed from strong to moderate expression in the kidney with ACE2 in the top 20% of the renal gene expression signal distribution (*Figure1A*). The analysis of human kidney single-cell data^28^ revealed a cell-type specific pattern of ACE2 expression, with proximal tubule cells showing the strongest expression signal (*Figure1B*). Our co-expression analysis conducted in both the discovery and replication data sets uncovered 47 kidney genes tightly correlated with ACE2 (*Figure1C*). Twenty-three (49%) of these genes showed expression specific to proximal tubule at the single-cell level (*Figure1C*). The ACE2-co-expressed genes were enriched for amino acid metabolism (P=1.89×10^-5^), mitochondria (P=1.92×10^-17^, *Figure1C*), kidney disease (P=1.88×10^-10^) and weakly for BP regulation (P=0.0225, *Figure1C*). Taken together, these data show that ACE2 is a highly abundant kidney gene with a cell-type specific pattern of expression in the proximal tubule and functional overlap with metabolic processes of key relevance to human health and disease.

**Figure1.**
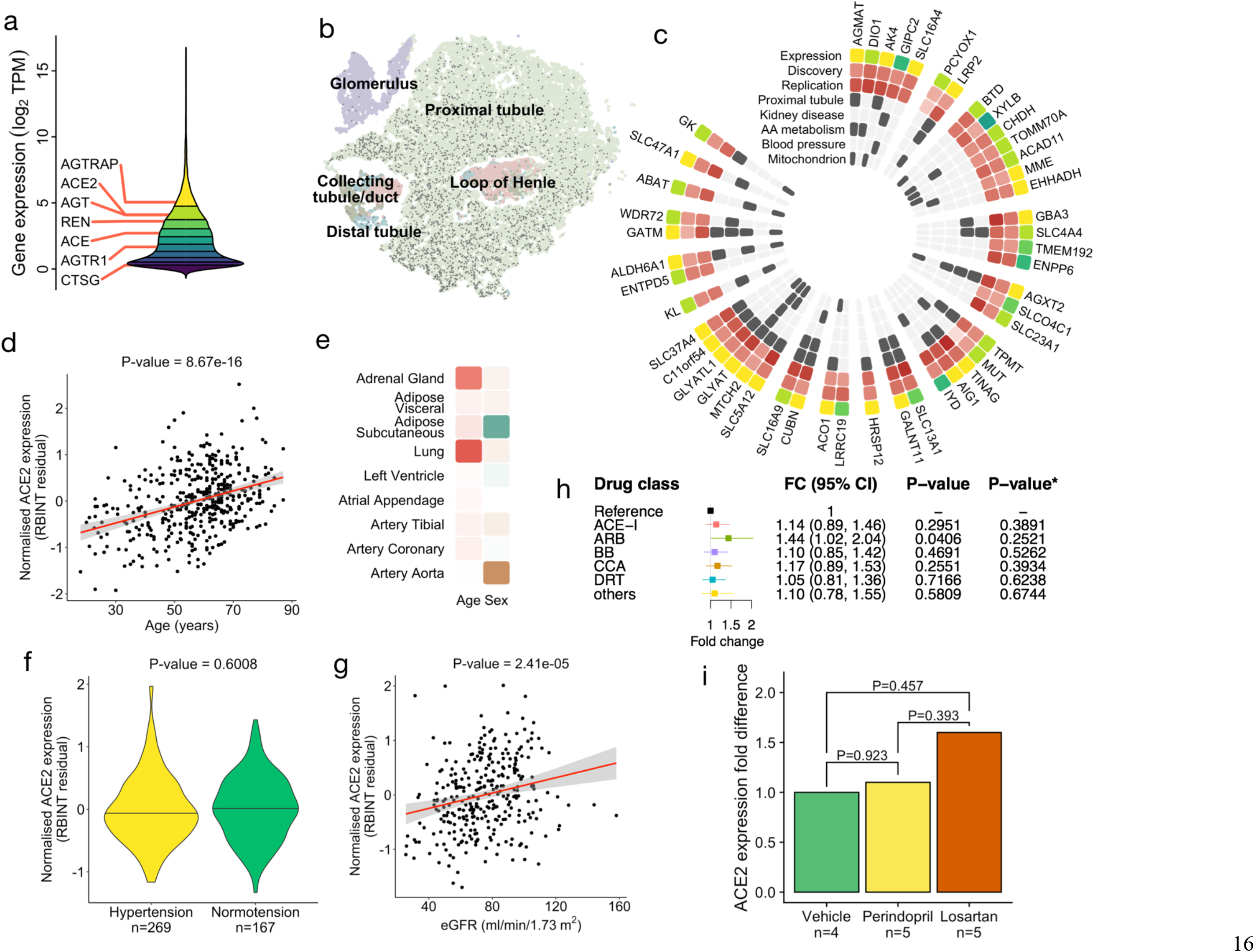
**a**. The distribution of gene expression values in the human kidney transcriptome derived from 436 samples of the discovery population. Deciles of the distribution are shown as coloured regions from least expressed decile (dark purple) to most highly expressed (yellow). Renal expression of the genes of the renin-angiotensin system are labelled (AGTRAP – angiotensin II receptor associated protein, ACE2 – angiotensin I converting enzyme 2, AGT – angiotensinogen, REN – renin, ACE – angiotensin I converting enzyme, AGTR1 – angiotensin II receptor type 1, CTSG – cathepsin G). Expression units are transformed (log2 of the sum of transcripts per million plus a constant offset of 1). **b**. Two-dimensional t-SNE representation of cells from normal kidney tissue. ACE2 expressing cells are marked with a dark grey circle. Cells are coloured by their location in the nephron segment. t-SNE – t-distributed stochastic neighbour embedding. **c**. Heatmap of 47 renal genes co-expressed with ACE2, genes are grouped and ordered by chromosomal location starting at the top and proceeding clockwise. Expression – quantile of expression in the distribution of the kidney transcriptome, colouring identical to A. Discovery – direction and strength of statistical association (t-statistic) in the discovery resource, positive association is shown from white (least strongly associated) to dark red (most strongly associated). Replication – direction and strength of statistical association (t-statistic) in the replication resource, colouring identical to “Discovery”. Proximal tubule – overlaps with the set of computational cell-type markers for proximal tubule cell – from Eales et al.^20^. Kidney disease – overlaps with “renal disease” set from genetic association database. AA-metabolism – present in REACTOME “Metabolism of amino acids and derivatives”. Blood pressure – present in the manually curated blood pressure genetically associated genes list. Mitochondrion – annotated with “G0:0005739” gene ontology term. **d**. Association between renal expression of ACE2 and age in the discovery data set. RBINT – the residual of normalised ACE2 expression, P-value – level of statistical significance. **e**. Heatmap of association between ACE2 expression and age and sex in the selected human tissues from Genotype-Tissue Expression project. The degree of positive association with age is coloured from white to dark red. A significant association with female sex is shown as green and with male sex as brown, less significant results scale towards white. **f**. Difference in renal expression of ACE2 between hypertensive and normotensive individuals in the discovery data set, n – number of individuals. **g**. Association between renal expression of ACE2 and estimated glomerular filtration rate (eGFR) in the discovery data set. **h**. The association between renal expression of ACE2 and each individual antihypertensive drug class, reference – hypertensive patients not on antihypertensive treatment, FC – fold change; 95% CI – 95% confidence interval; P-value – nominal P-value from the generalised estimation equation (gee) model; P-value* – adjusted P-value after correction for multiple testing. T vs N – treatment vs no treatment. **i**. Fold-change (calculated relative to “vehicle”) in renal ACE2 expression after 4-week treatment with perindopril and losartan in spontaneously hypertensive rats (SHR). P-values – level of statistical significance on ΔCt values from t-test (sample size shown).

### The effect of sex and age on ACE2 expression in the kidney and other human tissues

ACE2 is encoded by a gene on the short arm of the X chromosome (Xp22.2) and was reported to escape X chromosome inactivation (XCI) in some human tissues^30^. To examine whether there is the XCI-driven sex bias in the renal expression of ACE2, we compared male and female gene expression profiles from 436 kidneys within the discovery data set and 98 renal tissue samples from the replication resource. We first confirmed that the sex-specific X-chromosomal and Y-chromosomal gene controls (XIST and RPS4Y1, respectively) showed the expected sex-specific pattern of expression in kidneys from both gene expression data sets (*FigureS1A*). Our discovery analysis showed that women have approximately 1.36-fold higher expression of ACE2 in the kidney when compared to men (P=8.6×10^-9^) (*FigureS1B*). We then replicated this observation in an independent population – TCGA (P=8.86×10^-5^) (*FigureS1C*). In the combined analysis of 534 samples women had approximately 1.4-fold higher level of renal ACE2 expression when compared to men (*FigureS1D*). We then examined how sex influences ACE2 expression in the lung and several other relevant tissues from the GTEx project^38^. In some tissues (i.e. subcutaneous adipose tissue) ACE2 showed higher expression in women than men, while in others (i.e. aorta) there was the opposite pattern of sex-specificity in ACE2 expression (*Figure1E*).

Age showed a positive association with the renal expression of ACE2 in the discovery data set (8.67×10^-16^) (*Figure1D*). The directionality of the age-renal ACE2 relationship was consistent in the replication resource but the association did not reach the level of statistical significance (P=0.2540), possibly because of the smaller number of samples in TCGA. Similar to the findings in the kidney, we detected a statistically significant positive association between age and the expression of ACE2 in lungs from the GTEx (P=1.62×10^-4^, *FigureS1E*). In the majority of the examined human tissues the direction of the association between age and ACE2 expression was consistent with that observed in the kidney and the lungs (*Figure1E*).

Collectively, these data show that ACE2 exhibits a heterogeneous tissue-dependent sex-specific expression pattern in human tissues and that expression of ACE2 tends to increase with age in the kidney, lungs and the majority of tissues of relevance to cardiovascular system.

### Analysis of association between renal expression of ACE2, hypertension and other clinical phenotypes of potential relevance to COVID-19

Taking advantage of clinical information available in our discovery resource we examined whether hypertension and other comorbidities/phenotypes of potential relevance to COVID-19 (including diabetes, BMI, eGFR) were associated with renal expression of ACE2. Our analysis, conducted in 269 hypertensives and 167 normotensives, revealed no association between human hypertension and ACE2 expression in the kidney (P=0.6008) (*Figure1F, TableS2*). An additional sensitivity analysis restricted to 215 individuals who were not on antihypertensive treatment confirmed these findings (P=0.1212) (*FigureS1F, TableS2*). Neither diabetes nor BMI showed an association with kidney expression of ACE2 (P=0.8445 and P=0.8843, respectively) (*FigureS1G, TableS2*). However, we detected a significant positive association between renal expression of ACE2 and eGFR (P=2.41×10^-5^) (*Figure1G, TableS2*). We then replicated this finding through a meta-analysis of correlation between tubulointerstitial ACE2 expression and eGFR using 315 renal transcriptomes from an independent resource – Nephroseq (P=1.19×10^-17^) (*FigureS1I, TableS3*, www.nephroseq.org). Taken together, these data show that renal expression of ACE2 is positively associated with a biochemical index of kidney function but not with hypertension or other cardiovascular/metabolic comorbidities.

### The effect of antihypertensive medications on the renal expression of ACE2 – analysis of human and rat kidneys

Of 269 hypertensive individuals in our discovery tissue resource, 221 were on antihypertensive treatment. We first confirmed that antihypertensive therapy (as a simple binarised variable) was not associated with expression of ACE2 in the kidney (P=0.4176). We then allocated each prescribed antihypertensive medication into one of six categories consistent with the six main BP lowering drug classes, in 160 individuals with detailed information on their pharmacological therapy (*FigureS1H*). After replacing the binarised antihypertensive treatment indicator with the variables indicative of the six classes and after correction for inter-correlation between the drug classes, we re-examined if renal ACE2 expression was associated with individual antihypertensive classes. There was a nominally significant association with ARB but no significant associations after the correction for multiple testing (*Figure1H*). We then corroborated these results in SHR treated for 4 weeks with an ACE-I and an ARB for 4 weeks (*Figure1I*). Collectively, these data show that there is no association between commonly prescribed antihypertensive medications and renal expression of ACE2.

## DISCUSSION

Despite widespread speculation that hypertension (and especially drugs that inhibit RAS) would be associated with increased expression of ACE2 and that this is turn, lead to increased susceptibility to COVID-19, our study revealed no effect of either on the renal expression of ACE2. We also showed that while age and sex changed the abundance of ACE2 in human tissues, common metabolic comorbidities of hypertension, including diabetes and obesity index were not associated with renal expression of ACE2. Finally, we revealed enrichment for amino acid metabolism, mitochondria and kidney disease in the renal ACE2 co-expression analysis and demonstrated a positive association between renal ACE2 expression and a biochemical index of kidney function in the absence of the SARS-CoV-2 infection.

It is widely acknowledged that while viral pneumonitis is the main clinical manifestation of SARS-CoV-2 infection, COVID-19 is a multi-organ disease^43^ affecting the cardiovascular system, renal-urinary tract and other organs and tissues. The recent studies have revealed the tropism of SARS-CoV-2 to the renal epithelium^15^ and the post-mortem electron microscopy analyses demonstrated the presence of viral inclusion structures within the kidney^11,43,44^. The autopsy findings of patients who died of COVID-19 confirmed the prominent structural damage of tubular renal epithelium^45^ and it appears that the viral invasion of the kidney parenchyma may (at least to some extent) contribute to acute kidney injury in patients with COVID-19^46^. Importantly, ACE2 – a key trans-membrane receptor used by SARS-CoV-2 to gain entry to the host cells^2,11,47^ – is very abundant in the human kidney^48,49^, possibly more so than in the lungs^46^. These observations suggest that the kidney is one of the key target tissues for SARS-Cov-2.

Our kidney transcriptome profiling studies mapped a strong cell-specific ACE2 expression signal to the proximal tubule and showed a strong enrichment for this cell-type amongst the genes showing the highest level of renal co-expression with ACE2. The over-representation of genes that encode different enzymes and molecular transporters uncovered by our co-expression analysis appears consistent with the previously reported, but not widely acknowledged, role of ACE2 in metabolic regulation, i.e. acting as a chaperone for amino acid transport and metabolism^50^. We also observed strong (4.1x) enrichment for gene-products that localise in the mitochondria lending support for the hypothesis that viral disruption of mitochondrial oxygen sensing mechanisms may be relevant to SARS-CoV-2-driven hypoxia^51^. In addition, we reveal that many ACE2 co-expressed kidney genes show a well-established role in renal physiology and/or are strong determinants of renal health or the risk of kidney diseases. For example, LRP2 and CUBN encode megalin and cubilin – the receptors responsible for reabsorption of proteins, hormones, and vitamins in the proximal tubule and regulation of urinary excretion of protein^52^. Encoded by GALNT11, polypeptide N-Acetylgalactosaminyltransferase 11 acts a post-transcriptional modifier of LRP2, regulates its function as an endocytosis receptor and associates with proteinuria^53^. Klotho (a product of KL) operates as a multifunctional regulator of renal phosphate reabsorption, apoptosis, fibrosis, and cellular senescence; the renal depletion of KL plays a role in both acute kidney injury and chronic kidney disease^54,55^. SLC47A1 encodes a multidrug and toxin extrusion protein involved in the tubular secretion of creatinine and different drugs^56^. Several of the ACE2 co-expressed renal genes (including LRP2, CUBN, SLC47A1, GALNT1) were associated with eGFR, albuminuria and/or the risk of chronic kidney disease in the genome-wide association studies^57-59^. While the inactivation, loss-of-function and/or drop in renal expression of most of these molecules is generally interpreted as detrimental to kidney health^47,48,53^, in our analyses the expression of these genes showed a positive association with the renal abundance of ACE2. Together with the directionally consistent positive association between renal expression of ACE2 and eGFR in two independent datasets our data point to the nephro-protective function of ACE2.

We detected no association between renal expression of ACE2 and human hypertension in our analysis. We also observed only a very weak enrichment for BP regulation in our kidney ACE2 co-expression analysis. Thus, the renal expression of ACE2 is unlikely related to the reported over-representation of patients with elevated BP amongst those with COVID-19. Indeed, while almost half of Chinese^6^ and Italian^5^ patients diagnosed with COVID-19 were hypertensive and hypertension was more prevalent amongst those who died at intensive care units than in those who were discharged^5^ or those with more severe than non-severe COVID-19^6^, these associations were not adjusted for age – a key correlate of COVID-19-related morbidity and mortality. Being older, male sex, higher BMI, and diabetes have been linked to a higher risk of infection and/or adverse outcomes in patients with COVID-19^6^. Of these factors, only sex and age showed an association with renal ACE2 expression in our analyses. While the effect of sex on ACE2 showed a strong dependence on the type of tissue, the age-related increase in ACE2 was apparent in both the kidney and the lungs and directionally consistent in the other examined human tissues. Whether this age-driven increase in ACE2 represents a ubiquitous molecular mechanism enhancing the viral entry into the host cells or protecting them from a virus-mediated organ injury (through, for example, increased degradation of angiotensin II and/or synthesis of angiotensin 1-7) ^1,8,11,45,60,61^ remains to be established.

Our study provides an important new insight into how pharmacological blockade of RAS (a well-established strategy in the management of hypertension and other cardiovascular diseases) relates to ACE2 expression and its contemplated role in COVID-19-driven organ injury. Indeed, the RAS inhibitors (through their potential effects on ACE2 expression) have been considered as both potentially protective and harmful to patients infected by SARS-CoV-2. While ACE2 is not a direct molecular target of the RAS blockers, ACE-I reduce the levels of the ACE2 substrate (angiotensin II) while ARB block the angiotensin II-AT1 receptors interaction and thus have been proposed to attenuate the angiotensin II-mediated organ injury^60–64^. On the other hand, RAS blockers have been hypothesised to enhance the entry of SARS-CoV-2 into host cells and enhance the severity of organ damage in patients with COVID-19, possibly as a result of up-regulated ACE2 expression^7,8,62^. There is a widely acknowledged paucity of data linking ACE-I and ARB with ACE2 expression in the human tissues of relevance to COVID-19^60^. Indeed, the available evidence on how RAS blockers relate to ACE2 expression is based mostly on experimental models^61,65,66^ and clinical studies conducted using blood^66–70^. However, ACE2 operates mostly as a tissue enzyme^60,71^ and thus the findings from the previous studies cannot be extrapolated into the most relevant human tissues including human kidney. The demonstrated lack of association between the RAS blockers and renal expression of ACE2 shown in our study makes it unlikely that antihypertensive treatment driven up-regulation of this key functional SARS-CoV-2 receptor may alter individual risk of viral infection or influence clinical outcomes in COVID-19. These observations are further supported by the recent observational studies showing no association between treatment with RAS and the risk of infection^72^ and/or severe illness^73,74^. To this end, our data also lend support to the clinical consensus (endorsed by International Society of Hypertension, European Society of Hypertension and European Society of Cardiology) not to discontinue RAS blocker in patients with COVID-19^63^.

Our analysis has an obvious limitation inherent to the observational origin of the detected associations. However, our findings are based on the largest available discovery dataset with >400 human kidneys and wherever feasible – replicated in independent resources. We also provide data from other human organs as context to our findings and make use of both whole tissue transcriptome profiling and single-cell experiments.

In summary, we reveal findings of potential biological and epidemiological importance to the SARS-CoV-2 infection, i.e. the signature of age of ACE2 expression in tissues of relevance to COVID-19 and the lack of association between common cardiovascular and metabolic comorbidities of COVID-19 (i.e. hypertension, diabetes) on the renal expression of the SARS-CoV-2 entry receptor. Furthermore, the lack of association between renal expression of ACE2 and RAS blockers demonstrated in this study provides an important molecular argument in favour of safety of commonly prescribed BP lowering medications (RAS blockers) in patients with COVID-19. Finally, our data suggest that in the absence of SARS-CoV-2, renal ACE2 is positively associated with eGFR and lends support to the notion that renal ACE2 plays a more significant role in the local control of proximal tubule metabolism and kidney function rather than in systemic BP regulation^50^.

## Data Availability

Selective data are available upon request.

## ACKNOWLEDGMENTS

This work was supported by British Heart Foundation project grants PG/17/35/33001 and PG/19/16/34270 and Kidney Research UK grant RP_017_20180302 to MT, British Heart Foundation Personal Chair CH/13/2/30154 and Manchester Academic Health Science Centre: Tissue Bank Grant to BK, Medical University of Silesia grants KNW-1-152/N/7/K to JZ and KNW-1-171/N/6/K to WW. LMB acknowledges support from National Health and Medical Research Council Program Grant (APP1055214) and Medical Research Future Fund (APP 1175865). Access to TCGA kidneys and GTEx data has been granted by NIH (approvals 50804-2 and 50805-2).

